# Impact of social distancing measures on the daily number of new COVID-19 cases in Côte d’Ivoire: a retrospective cohort study

**DOI:** 10.1101/2021.04.18.21255693

**Authors:** Teegwende V. Porgo, Khadidja Malloum Boukar, Ezechiel A. Djallo, Richard Quansah Amissah, Coralie Assy, Adama Traore, Gaston Sorgho

## Abstract

**Introduction:** Côte d’Ivoire is facing a second wave of the novel coronavirus disease 2019 (COVID-19). While social distancing measures (SDM) may be an option to address this wave, SDM may be devastating, especially if they have a minimal impact on the spread of COVID-19, given the other measures in place.

**Methods:** We conducted a cohort study involving cases that had occurred as at June 30, 2020. We used data from the Government’s situation reports. We established three study periods, which correspond to the implementation and easing of SDM, including a 10-day delay for test results: (1) the SDM (March 11 - May 24), (2) the no SDM (May 25 - June 21), and (3) the pseudo SDM (June 22 - July 10) periods. We compared the incidence rate during these periods using Poisson regression, with sex, age, and the average daily number of tests as covariates.

**Results:** As at July 10, there were 12,052 cases. The incidence rate was 100% higher during period 2 compared to period 1 (incidence rate ratio = 2.05, 95% confidence interval: 1.75-2.41) and 25% lower during period 3 compared to period 2 (0.75 [0.66-0.86]).

**Conclusions:** The easing and subsequent reinforcement of SDM had a significant impact on the spread of COVID-19 in Côte d’Ivoire. The other mitigation measures either did not compensate for the easing of the SDM during the no SDM period or were not fully effective throughout the study periods; they should be strengthened before the SDM are reimplemented.

## Introduction

The first case of the novel coronavirus disease 2019 (COVID-19) was confirmed in Côte d’Ivoire on March 11, 2020.^1^ In the beginning of the pandemic, the Government focused its COVID-19 emergency preparedness and response plan on preventive measures, including hygiene and social distancing measures (SDM), and the gradual strengthening of laboratory testing and medical surge capacity.

After the peak of the first wave of COVID-19 in the country, several preventive measures were lifted and the reinforcement of other measures, such as mandatory mask wearing, ceased; however, the number of new COVID-19 cases has been increasing steadily since mid-December 2020.^1^ While other countries have faced their second and/or third waves by re-imposing SDM, such measures could be devastating in a lower middle-income country like Côte d’Ivoire. This would be all the more dramatic if SDM have a minimal impact on the spread of COVID-19, given the other measures in place. The objective of this study was to assess the impact of SDM on the daily number of new cases of COVID-19 in Côte d’Ivoire.

## Methods

### Study design and population

We conducted a retrospective cohort study involving all COVID-19 cases that occurred in Côte d’Ivoire between March 11, 2020 and June 30, 2020. A COVID-19 case was defined as an individual who tested positive for the novel coronavirus 2019 following a real-time reverse transcriptase polymerase chain reaction test.

Different SDM were implemented in the greater Abidjan (the economic capital city) region and in the rest of the country (Figure 1).^2-7^ We focused on measures that were implemented in the greater Abidjan region because: (1) between March 11, 2020 and June 30, 2020, 95.44% of all COVID-19 cases resided in that region and (2) that region was isolated from the rest of the country between March 29, 2020 and July 15, 2020. These measures, which included the implementation of a curfew; closure of restaurants, schools, and universities; and limiting social gatherings to a maximum of 50 people (including gatherings in places of worship), were implemented between March 16, 2020 and March 24, 2020. Effective May 15, 2020, the curfew was lifted nation-wide, restaurants were permitted to reopen, and the maximum number of people that could gather was increased to 200. Schools and universities begun reopening on May 25, 2020. Faced with an upsurge in cases in Mid-May 2020, the Government reduced the number of people that could gather to 50 people (including gatherings in restaurants and places of worship) on June 12, 2020. All this while, bars, night clubs, cinemas, and other entertainment venues were closed until July 31, 2020. Consequently, we considered three periods for our study: (1) the SDM period (March 11 - May 14), (2) the no SDM period (May 15 - June 11), and (3) the pseudo SDM period (June 12 - June 30). Since test results were available with a delay of 3 days at the very beginning of the pandemic and 7-10 days subsequently, we applied a 10-day delay to these periods: March 11 - May 24, May 25 - June 21, and June 22 - July 10.^7^

**Figure 1.**
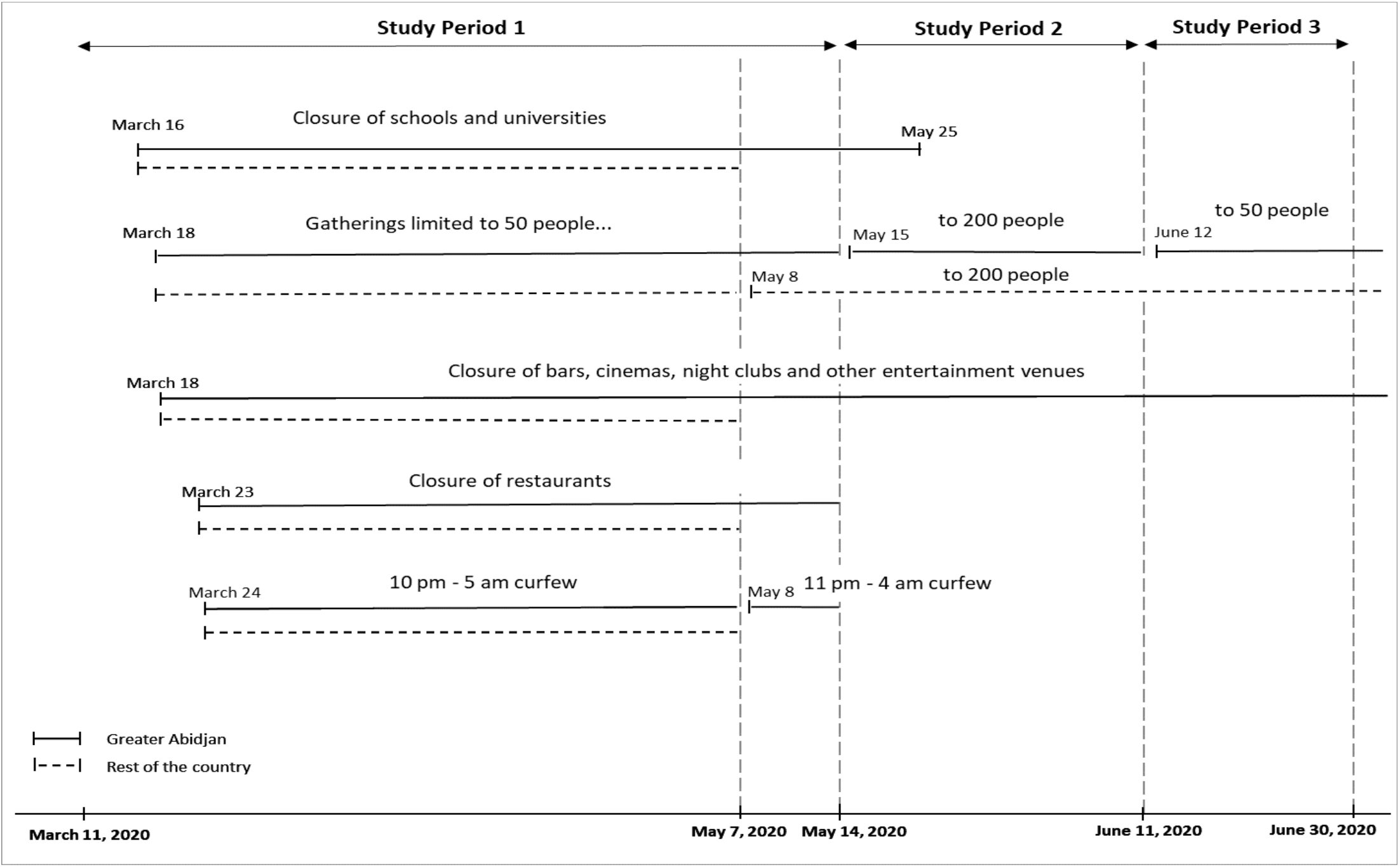
Social distancing measures implemented in Côte d’Ivoire^2-7^.

### Study data

We used data from the COVID-19 situation reports produced by the Government with the support of the World Health Organization.^1^ Each report included aggregated, cumulative data as at the day before the date of publication of the report. The data included the number of cases (including the number of deaths), the positive test rate, and the distribution of cases by region, health district, sex, and age categories.

### Statistical Analyses

Using Poisson regression with a robust estimator of the variance, we compared the average daily number of new COVID-19 cases (incidence rate) during the different study periods. First, we checked for model assumptions; that is, we confirmed that the incidence rate of COVID-19 followed a Poisson distribution and that the log of the mean rate was a linear function of the daily number of tests. In each model, we included the variables sex, age (0-10, 11-20, 21-30, 31-40, 41-50, ≥51 years), and average daily number of laboratory tests as covariates and checked for collinearity. The average daily number of tests was calculated using the positive test rate and included in the model as the “exposition variable.”^8^ Data on age were missing for 6.80% of cases. We imputed these data with the mode age; the mode age among men was the same as that among women (31-40 years). We calculated incidence rate ratios with 95% confidence intervals.

We performed statistical analyses using the STATA software (StataCorp LLC, College Station, TX, version 15.1). Statistical tests were two-sided with α = 0.05.

## Results

As at July 10, 2020, 12,052 COVID-19 cases were recorded in Côte d’Ivoire. Of these, 7,114 (59.03%) were males and 1,403 (11.64%), 3,504 (29.07%), and 3,757 (31.17%) resided in Yopougon Est, Treichville-Marcory, and Cocody-Bingerville, respectively. The cumulative number of deaths was 81, of whom 45 (55.56%) were ≥61 years old and 60 (74.07%) were males. The average daily number of tests during the SDM, no SDM, and pseudo SDM periods was 288, 803, and 1,412, respectively.

Compared to the SDM period, the incidence rate of COVID-19 was 100% higher in the no SDM period (incidence rate ratio = 2.05 [1.75-2.41]) and 54% higher in the pseudo SDM period (1.54 [1.31-1.82]; Table 1). Compared to the no SDM period, the incidence rate of COVID-19 was 25% lower in the pseudo SDM period (0.75 [0.66-0.86]; model results not shown). The incidence rate of COVID-19 in individuals in the 31-40 years age group was 27 times that in children who were 10 years old or less (26.89 [19.88-36.37]).

**Table 1.** Incidence rate ratios of the daily number of new COVID-19 cases based on the implemented social distancing measures at the time.

| Characteristics | N (%) | Incidence rate ratios [95% CI] |
| --- | --- | --- |
| **Overall** | 12,052 (100) |  |
| **Sex** |  |  |
| Female | 4938 (40.97) | - |
| Male | 7114 (59.03) | 1.39 [1.25-1.55] |
| **Age (years)** |  |  |
| 0-10 | 191 (1.58) | - |
| 11-20 | 597 (4.95) | 3.67 [2.49-5.41] |
| 21-30 | 2357 (19.56) | 14.33 [10.48-19.61] |
| 31-40 | 4340 (36.01) | 26.89 [19.88-36.37] |
| 41-50 | 2521 (20.92) | 15.56 [11.41-21.22] |
| ≥ 51 | 2046 (16.98) | 12.22 [8.64-17.29] |
| **Test\*** | 70,894 (100) | 1 |
| **Period** |  |  |
| 1 | 2376 (19.71) | - |
| 2 | 5116 (42.45) | 2.05 [1.75-2.41] |
| 3 | 4560 (37.84) | 1.54 [1.31-1.82] |

N: number of cases or tests, CI: confidence interval, period 1, 2 and 3: the social distancing, no social distancing and pseudo social distancing periods, respectively
\*The average daily number of tests was included in the model as “the exposition variable”<sup>8</sup>

## Discussion

In this retrospective cohort study, we examined the impact of SDM on the incidence of COVID-19 in Côte d’Ivoire between March 11 and June 30; most cases (69.08%) resided in Yopougon Est, Treichville-Marcory, and Cocody-Bingerville. The measures included the implementation of a curfew, the closure of restaurants, schools, and universities, and restrictions on gatherings (including gatherings in places of worship). When the Government lifted the curfew, allowed restaurants, schools, and universities to reopen, and eased restrictions on gathering, the incidence rate increased by 100%. Subsequently, when the Government reinforced gathering restrictions, the incidence rate decreased by 25%. The incidence rate was higher in the 31-40 years age group.

Thus far, it was believed that the upsurge in cases in Côte d’Ivoire during the first wave was due to the strengthening of the country’s testing capacity.^9^ This is understandable since when the daily number of tests was omitted from the analyses, the incidence rate increased by 573% during the no SDM period, compared to the SDM period, and by 32% during the pseudo SDM period, compared to the no pseudo SDM period (results not shown). However, our study (in which we adjusted for sex, age, and testing capacity) showed that this upsurge was due, at least in part, to the lifting of SDM.

Breaking the chain of transmission of a virus requires the curtailing of (i) interactions between infected and uninfected people, (ii) infectiousness of infected people, and/or (iii) susceptibility of uninfected people.^10^ In the absence of antivirals specific to COVID-19 and limited vaccines availability, curtailing interactions between infected and uninfected people is the only option to fight COVID-19. This option involves two strategies: (i) SDM and (ii) policies of screening, isolation of infected people, and quarantining contacts of infected people.^10,11^

SDM and policies of screening, isolation, and quarantine are additive such that relaxing efforts towards one strategy should necessarily be accompanied by the reinforcement of the other strategy. Moreover, SDM are not “a necessary cause,” in a sense that, if we can implement mass or strategic screening and ensure that cases are isolated and contacts are quarantined for at least 14 days (the equivalent of the COVID-19 incubation period), this in itself can reduce the number of cases and bring the epidemic under control.^12,13^ Mass or strategic screening is important because SDM are not a sustainable solution. They significantly disrupt health and education systems, increase social inequality, slow economic growth, and can reverse development gains, all of which ultimately impact health outcomes.^14-19^ For example, disruption of health systems and decreased access to food could increase the monthly child deaths by 9.80% (less severe scenario) or 44.70% (most severe scenario) and the monthly maternal deaths by 8.30% or 38.60%.^14^ Furthermore, when COVID-19 vaccines become available in Côte d’Ivoire, the risk of infection with the coronavirus 2019 would not be eliminated quickly since the country would first need to strengthen its vaccine deployment capacity, which would include conducting effective awareness campaigns and improving the country’s cold chain and strategies for last-mile delivery to rural areas.^10,20,21^

This study suggests that SDM had a significant impact on the spread of COVID-19 in Côte d’Ivoire, as has been observed in several other countries.^22^ The study also suggests that the screening, isolation, and quarantining strategies that were implemented either did not compensate for the easing of the SDM during the no SDM period or were not fully effective during all the three study periods. Either way, the low number of daily tests, the difficulty of ensuring that cases that are not treated in health facilities are self-isolating, and the difficulty of tracing and quarantining contacts during the study periods should be remedied moving forward.

As of January 28, 2021, the average daily number of tests in the country was approximately 1020, which is low when compared to the size of the country (more than 26 million) or even the city Abidjan (more than 4 million).^1,23,24^ Moreover, on June 25, the Government announced that test results would be made available within 48 hours; however, this needs to be reinforced.^25^ Since up to 88% of cases may be asymptomatic and these cases are infectious, it would be beneficial to move from the current voluntary testing scheme to a more active one.^26-28^ Given the limited resources of the country, mass testing at the national level (or regional level) may not be an appropriate option. However, strategic mass testing could be implemented, targeting individuals who are more likely to be infected, for example high-risk age groups in the most affected neighborhoods. Moreover, saliva-based tests could enable mass testing amid scarce testing resources. Saliva-based tests are cheaper and less invasive than the other testing tools currently available.^29,30^ Furthermore, most cases in Côte d’Ivoire are not treated in healthcare facilities. These cases need to be closely monitored to ensure self-isolation for 14 days.

Contact tracing is also critical, especially since a high proportion of COVID-19 cases may be asymptomatic and, in Côte d’Ivoire, as of the writing of this paper in February 2021, contacts that are identified are not systematically tested if they do not have any symptoms. The average number of new infections generated by an infectious person in a fully susceptible population has been estimated to be 2.37 in Africa.^31^ Moreover, data on natural immunity are mixed.^32,33^ Consequently, it can be considered that each of the 12,052 cases included in this study infected 2 contacts, resulting in 24,104 infected contacts. Yet, as of the end of our study period, 12,408 contacts had been identified, which implies that at least 11,696 were missed by the contact tracing system. In this regard, Côte d’Ivoire could greatly benefit from emulating Vietnam, another lower middle-income country. Although the first COVID-19 case was reported in Vietnam on January 23, the country had 1657 COVID-19 cases (and 35 deaths) as at January 29, 2021.^34^ The spread of the virus in Vietnam was reduced in households and in the community thanks, in part, to the country’s effective contact tracing strategy which identifies contacts of cases and the subsequent contacts of contacts of cases.^35^ First, contacts are identified through the collaborative effort of health professionals, law-enforcement officers, and other government officers. These individuals aim to identify any individual who might have been in close contact with a case in the previous 14 days. Second, contacts are tested; contacts who test positive are isolated in a health care facility while those who test negative are quarantined at designated centers for 14 days. Third, contacts of contacts must self-isolate for 14 days.

### Limits

Since there were short intervals of time between the implementation (and easing or lifting) of SDM, we could not assess the impact of each SDM. Therefore, we compared “periods of SDM.” Additionally, test results were not readily available, and the delays varied. Thus, we distinguished between the time of occurrence and the time of reporting of cases and applied the maximum delay in test results to each study period.

## Conclusions

The easing and subsequent reinforcement of SDM had a significant impact on the incidence rate of COVID-19 in Côte d’Ivoire between March and June 2020. Our results suggest that the other mitigation measures either did not compensate for the easing of the SDM during the no SDM period or were not fully effective throughout the study periods. These measures should be reinforced before SDM are reimplemented.

## Data Availability

The data analysed in the study is available upon request.

The authors did not receive any funding for this study.

The authors declare that they have no competing interests.

The findings and conclusions in this article are those of the authors and do not necessarily represent the official position of the World Bank Group or the Republic of Côte d’Ivoire.

## Authors’ contributions

Study conception: TVP

Study design: TVP, KM, ED

Data acquisition: AT, TVP, GS

Data analyses: ED, KM, TVP

Data interpretation: all authors

Drafting of manuscript: TVP, RQA, CA

Manuscript critical revision for intellectual content: all authors

